# Estrogen and COVID-19 symptoms: associations in women from the COVID Symptom Study

**DOI:** 10.1101/2020.07.30.20164921

**Authors:** Ricardo Costeira, Karla A. Lee, Benjamin Murray, Colette Christiansen, Juan Castillo-Fernandez, Mary Ni Lochlainn, Joan Capdevila Pujol, Heather Macfarlane, Louise C. Kenny, Iain Buchan, Jonathon Wolf, Janice Rymer, Sebastien Ourselin, Claire J. Steves, Timothy D. Spector, Louise R. Newson, Jordana T. Bell

**Affiliations:** Department of Twin Research and Genetic Epidemiology, King’s College London, London, U.K; School of Biomedical Engineering & Imaging Sciences, King’s College London, London, U.K; Zoe Global Limited, London, U.K; National Health Service, U.K; Department of Women’s and Children’s Health, University of Liverpool, U.K; Department of Public Health, Policy and Systems, University of Liverpool, U.K; Department of Women’s Health, King’s College London, London, U.K; Newson Health Menopause & Wellbeing Centre, Stratford-Upon-Avon, U.K

## Abstract

**Background:** Men and older women have been shown to be at higher risk of adverse COVID-19 outcomes. Animal model studies of SARS-CoV and MERS suggest that the age and sex difference in COVID-19 symptom severity may be due to a protective effect of the female sex hormone estrogen. Females have shown an ability to mount a stronger immune response to a variety of viral infections because of more robust humoral and cellular immune responses.

**Objectives:** We sought to determine whether COVID-19 positivity increases in women entering menopause. We also aimed to identify whether premenopausal women taking exogenous hormones in the form of the combined oral contraceptive pill (COCP) and post-menopausal women taking hormone replacement therapy (HRT) have lower predicted rates of COVID-19, using our published symptom-based model.

**Design:** The COVID Symptom Study developed by King’s College London and Zoe Global Limited was launched in the UK on 24^th^ March 2020. It captured self-reported information related to COVID-19 symptoms. Data used for this study included records collected between 7^th^ May - 15^th^ June 2020.

**Main outcome measures:** We investigated links between COVID-19 rates and 1) menopausal status, 2) COCP use and 3) HRT use, using symptom-based *predicted* COVID-19, tested COVID-19, and disease severity based on requirement for hospital attendance or respiratory support.

**Participants:** Female users of the COVID Symptom Tracker Application in the UK, including 152,637 women for menopause status, 295,689 for COCP use, and 151,193 for HRT use. Analyses were adjusted for age, smoking and BMI.

**Results:** Post-menopausal women aged 40-60 years had a higher rate of *predicted* COVID (P=0.003) and a corresponding range of symptoms, with consistent, but not significant trends observed for tested COVID-19 and disease severity. Women aged 18-45 years taking COCP had a significantly lower *predicted* COVID-19 (P=8.03E-05), with a reduction in hospital attendance (P=0.023). Post-menopausal women using HRT or hormonal therapies did not exhibit consistent associations, including increased rates of *predicted* COVID-19 (P=2.22E-05) for HRT users alone.

**Conclusions:** Our findings support a protective effect of estrogen on COVID-19, based on positive association between *predicted* COVID-19 and menopausal status, and a negative association with COCP use. HRT use was positively associated with COVID-19 symptoms; however, the results should be considered with caution due to lack of data on HRT type, route of administration, duration of treatment, and potential comorbidities.

**Trial registration:** The App Ethics has been approved by KCL ethics Committee REMAS ID 18210, review reference LRs-19/20-18210

## Introduction

As the COVID-19 pandemic progresses, it has been widely observed that adult men of all ages are at higher risk of developing serious complications. A recent review of biological sex and COVID-19 has described the male bias in COVID-19 mortality in 37 of the 38 countries that have provided sex-disaggregated data (1). Of women who develop COVID-19, being post-menopausal has been independently associated with more severe COVID-19 (2). Epidemiological data from previous coronavirus outbreaks, including SARS-CoV (Severe Acute Respiratory Syndrome Corona Virus) and MERS-CoV (Middle East Respiratory Syndrome Corona Virus) showed the same pattern: among men, morbidity and fatality rates were markedly higher compared to women (3, 4). When patients aged 70 years or older were examined, the effect of lower morbidity and mortality disappeared among SARS-CoV-2 infected women (5). It has also been noted that pregnant women experience milder COVID-19 than initially expected (6).

Animal model studies of SARS-CoV and MERS suggest that the age and sex difference in COVID-19 symptom severity may be due to protective and acute actions of the female sex hormone estrogen (7). Females have been shown to be able to mount a stronger immune response to a variety of viral infections because of more robust humoral and cellular immune responses (8-10). Anti-mullerian hormone (AMH) and estradiol are markers of high ovarian reserve and have been shown to negatively correlate with severity of COVID-19, independent of age, suggesting that pre-menopausal women are somewhat protected against more severe COVID-19 (11). A phase II study testing whether short-course estradiol delivered via transdermal patch will be safe and reduce symptom severity in COVID-19 affected adult men and older women is underway in New York (NCT04359329), with another study investigating oral progesterone in men hospitalised with COVID-19 in California (NCT04365127).

The potential protective effect of estrogen against COVID-19 requires continued and careful evaluation. Here, we investigate whether higher levels of estrogen are linked to a reduction in the rate and severity of COVID-19 among women, based on large-scale self-reported data from the UK. The primary analysis explores if women who have recently gone through menopause are at higher risk of symptomatic COVID-19, compared to women who still report periods over age 40. Follow-up analyses consider exogenous estrogen, exploring if women taking the combined oral contraceptive pill (COCP) have a reduced risk of being COVID-19-positive, with associated symptoms. We also assess if use of HRT is associated with a reduced rate of COVID-19 positivity and severity in post-menopausal women alone.

## Materials and Methods

The COVID Symptom Study Smartphone Application ("app") was developed by Zoe Global Limited with scientific input from researchers and clinicians at King’s College London and Massachusetts General Hospital. It was launched in the UK on 24^th^ March 2020. It captures self-reported information related to COVID-19 symptoms. On first use, the app records self-reported location, age, and core health risk factors. At this point height and weight are self-reported, allowing calculation of body mass index (BMI). With continued use, participants provide daily updates on symptoms, information on health care visits, COVID-19 testing results, and whether they are self-quarantining or seeking healthcare, including the level of intervention and related outcomes. Individuals without apparent symptoms are also encouraged to use the app.

On 7 May 2020 we asked all female app-users if they are presently taking any forms of hormonal therapies including hormone replacement therapy (HRT), hormonal contraceptives and testosterone (**S1 Fig**). We also posed questions relating to menstruation and current pregnancy. Patients who indicated they were still having periods were asked about frequency of menstruation. Those who indicated not having periods were asked their age at menopause. The COVID Symptom Study dataset used for this study was obtained from the period of 7 May - 15 June 2020, yielding 40 days of data collection from a total of 1.9M women in the UK. From these, 1.6M women had BMI between 20-35kg/m^2^ and were included in downstream analyses.

### Ascertainment of exposures, disease outcomes and study covariates

Exposures, outcomes and covariates were ascertained from self-reported app data following quality control with purpose-built scripts (https://github.com/KCL-BMEIS/zoe-data-prep). Exposures used in our analyses included women’s menopausal status, and COCP and HRT use. Primary disease outcomes included COVID-19-related symptoms (**S1 Table**) and *predicted* COVID-19 positivity based on symptoms, as described in our recent publication (12). Hospitalisation and respiratory support, defined as supplementary oxygen (+/- ventilation) were used as surrogate markers for disease severity. Self-reported results from nose/throat swab tests were also used as outcome for a subset of the sample who were tested for COVID-19. Reported age, BMI and smoking status were used as covariates in the associations, and the analyses only included female app users.

Analyses of menopausal status included post- and pre-menopausal women aged 40-60 years, with BMI 20–35 kg/m^2^, excluding women taking any form of hormonal therapy. We compared post-menopausal women currently reporting no periods and with last period reported after the age of 40 and within 5 years, to pre-menopausal women with regular periods occurring every 3–6 weeks. Analyses of COCP use included post- and premenopausal women aged 20-45 years, with BMI 20–35kg/m^2^. We compared women taking COCP as their only form of hormonal therapy, to women of the same age taking no form of hormone therapy. Analyses of HRT use were carried out in post-menopausal women aged 50-65 years, with BMI 20–35kg/m^2^ and last periods reported at age 45–60. We compared women on HRT alone to women receiving no form of hormonal therapy. Extended hormone therapy analyses also considered women who were on HRT or related hormone therapies, including COCP, progestogen therapy, progestogen containing intrauterine systems, or testosterone (**S1 Table**). Use of estrogen for gender transitioning was excluded from the analyses.

### Association analyses

Binomial generalized mixed models with a log-odds/logit link function were used to carry out association analyses. The models included COVID-19 symptom or outcome as a function of exposure variables and covariates including age, BMI, and smoking status. Exposures included menopausal status, COCP use, and HRT use. Exposure data and symptoms of the disease were coded as ‘1’ for positive (TRUE/yes/severe/significant) responses and ‘0’ for negative (FALSE/no/mild) responses or blank (NA) statements from app users. In *predicted* COVID-19 analyses subjects with a *predicted* COVID-19 probability ≥ 50% were considered COVID-19-positive (coded ‘1’, while a predicted COVID-19-negative outcome was coded as ‘0’). App users in hospital or home from hospital were coded as ‘1’ for hospitalisation. App users who reported going to the hospital and requiring supplementary oxygen (+/-ventilation) were coded as ‘1’ for respiratory support, while all others were coded as ‘0’ for this outcome. App users with at least one positive swab test for COVID-19 were considered COVID-19-positive in the *tested* COVID-19-positive analyses (coded as ‘1’, while a tested COVID-19-negative outcome was coded as ‘0’). Only a small subset of women was tested for COVID-19, and missing data, failed tests, or tests awaiting outcome were excluded from analyses. Age and BMI were coded as continuous fixed effects. Only women with BMI 20–35kg/m^2^ were considered in our study. Smoking was coded as a categorical fixed effect variable with levels ‘1’, ‘2’ and ‘3’ respectively representing never-smokers, ex-smokers and current smokers. Odds ratios of the associations are reported.

### Sensitivity analyses

Age is a key risk factor for COVID-19. Age sensitivity analyses were performed to match the mean and median ages of cases and controls for each of the three exposure variables. Analyses were carried out in subsets of app users within 5-year bins for menopausal status and use of COCP and HRT. In menopause, sensitivity analysis was also performed for years passed since time of last period.

### TwinsUK cohort data analysis

At the time of data freeze, a subset of app users included research volunteers from the TwinsUK cohort (13). Here, we considered 270 TwinsUK female twins (mean age 66), who also had existing whole blood DNA methylation data profiled using the Illumina Infinium HumanMethylation450 BeadChip. From the 270 female twins, a subset of 84 participants had previously collected TwinsUK questionnaire data on menopausal status, and a further subset of 75 had information on age at last period.

Whole blood DNA extraction and DNA methylation profiling in these samples have been previously described (14). Briefly, DNA methylation levels were determined using Illumina methylation beta-values (15), which range between 0 at unmethylated CpG-sites, and 1 at fully methylated CpG sites. ENmix (16) was used for quality control and minfi (17) was used to exclude samples with median methylated and unmethylated signals below 10.5. Three different epigenetic age calculators were applied to estimate DNA methylation age for each individual, DNAm GrimAge (18), DNAm PhenoAge (19) and the original Horvath methylation age (20). Epigenetic age acceleration measures included estimates obtained from regressing epigenetic age on chronological age. We also considered intrinsic epigenetic age acceleration (IEAA) and extrinsic epigenetic age acceleration (EEAA) (21). IEAA is calculated by regressing the Horvath DNA methylation age and cell blood counts to create an estimate of cell intrinsic methylation ageing, independent of differences in blood cell counts. EEAA is calculated based on epigenetic ageing measures developed by Hannum et al. (22), up-weighted by the relative proportion of three age-related blood immune cells, specifically naïve (CD45RA+CCR7+) cytotoxic T cells, exhausted (CD28-CD45RA-) cytotoxic T cells, and plasmablasts, as previously described (21). EEAA has been proposed to capture aspects of age-related immuno-senescence (21). A one-sided t-test was applied to compare each of the five age acceleration measures to symptoms and *predicted* COVID-19 status, and results are presented at nominal significance.

## Results

The COVID Symptom Study dataset used for this study was collected between 7 May - 15 June 2020 and included 1.6M women in the UK (BMI range 20–35kg/m^2^), with all analyses adjusted for age, BMI and smoking status.

### Menopause

We examined the impact of entering menopause on COVID-19-positivity and related symptoms among 152,637 women aged 40–60 years with BMI 20-35 kg/m^2^. Cases were defined as post-menopausal women currently reporting no periods and with last period reported after the age of 40 and within the last 5 years, resulting in altogether 44,268 women (**S1 Table**). Controls were defined as pre-menopausal women with regular periods occurring every 3–6 weeks, resulting in altogether 108,369 controls. Women taking any form of hormonal therapy were excluded from this analysis.

Post-menopausal women had a higher rate of *predicted* COVID-19 (OR = 1.22, 95% CI 1.07–1.39, *p* = 0.003) and a corresponding range of significant differences in symptoms including hoarse voice, skipped meals, muscle pains, and fever (**Table 1**). Requirement for hospitalisation and respiratory support were not significant, but also showed a positive direction of association in post-menopausal women. Although there was no significant association between menopausal status and testing COVID-19-positive, the direction of association is consistent with *predicted* COVID-19 results (**Table 1**).

**Table 1.**
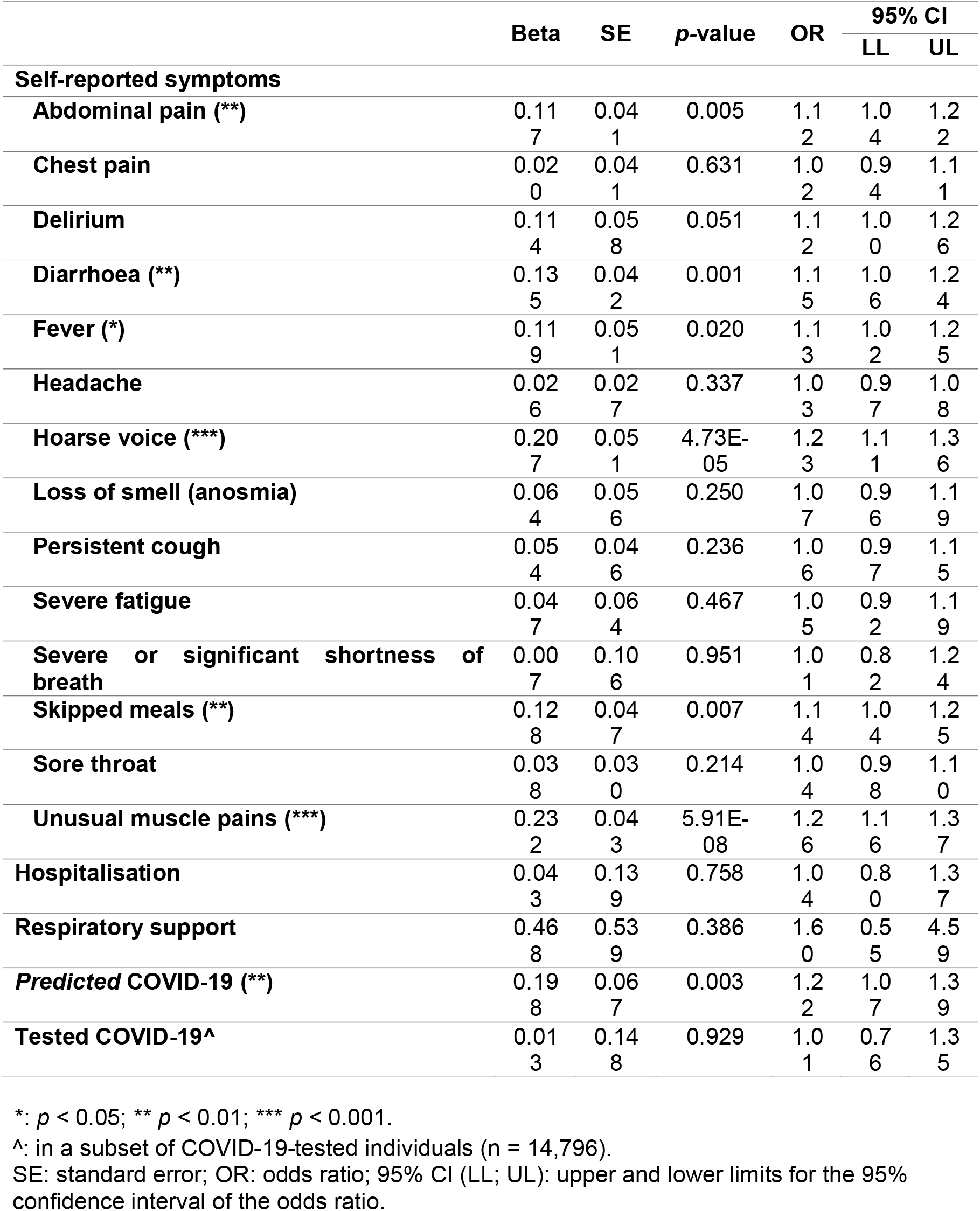
Association of menopausal status to *predicted* and tested COVID-19 results, self-reported symptoms and indicators of COVID-19 disease severity (hospitalisation/respiratory support) (n = 152,637).

|  | Beta | SE | p-value | OR | 95% CI |  |
| --- | --- | --- | --- | --- | --- | --- |
|  |  |  |  |  | LL | UL |
| **Self-reported symptoms** |  |  |  |  |  |  |
| **Abdominal pain (\*\*)** | 0.11<br>7 | 0.04<br>1 | 0.005 | 1.1<br>2 | 1.0<br>4 | 1.2<br>2 |
| **Chest pain** | 0.02<br>0 | 0.04<br>1 | 0.631 | 1.0<br>2 | 0.9<br>4 | 1.1<br>1 |
| **Delirium** | 0.11<br>4 | 0.05<br>8 | 0.051 | 1.1<br>2 | 1.0<br>0 | 1.2<br>6 |
| **Diarrhoea (\*\*)** | 0.13<br>5 | 0.04<br>2 | 0.001 | 1.1<br>5 | 1.0<br>6 | 1.2<br>4 |
| **Fever (\*)** | 0.11<br>9 | 0.05<br>1 | 0.020 | 1.1<br>3 | 1.0<br>2 | 1.2<br>5 |
| **Headache** | 0.02<br>6 | 0.02<br>7 | 0.337 | 1.0<br>3 | 0.9<br>7 | 1.0<br>8 |
| **Hoarse voice (\*\*\*)** | 0.20<br>7 | 0.05<br>1 | 4.73E-05 | 1.2<br>3 | 1.1<br>1 | 1.3<br>6 |
| **Loss of smell (anosmia)** | 0.06<br>4 | 0.05<br>6 | 0.250 | 1.0<br>7 | 0.9<br>6 | 1.1<br>9 |
| **Persistent cough** | 0.05<br>4 | 0.04<br>6 | 0.236 | 1.0<br>6 | 0.9<br>7 | 1.1<br>5 |
| **Severe fatigue** | 0.04<br>7 | 0.06<br>4 | 0.467 | 1.0<br>5 | 0.9<br>2 | 1.1<br>9 |
| **Severe or significant shortness of breath** | 0.00<br>7 | 0.10<br>6 | 0.951 | 1.0<br>1 | 0.8<br>2 | 1.2<br>4 |
| **Skipped meals (\*\*)** | 0.12<br>8 | 0.04<br>7 | 0.007 | 1.1<br>4 | 1.0<br>4 | 1.2<br>5 |
| **Sore throat** | 0.03<br>8 | 0.03<br>0 | 0.214 | 1.0<br>4 | 0.9<br>8 | 1.1<br>0 |
| **Unusual muscle pains (\*\*\*)** | 0.23<br>2 | 0.04<br>3 | 5.91E-08 | 1.2<br>6 | 1.1<br>6 | 1.3<br>7 |
| **Hospitalisation** | 0.04<br>3 | 0.13<br>9 | 0.758 | 1.0<br>4 | 0.8<br>0 | 1.3<br>7 |
| **Respiratory support** | 0.46<br>8 | 0.53<br>9 | 0.386 | 1.6<br>0 | 0.5<br>5 | 4.5<br>9 |
| **Predicted COVID-19 (\*\*)** | 0.19<br>8 | 0.06<br>7 | 0.003 | 1.2<br>2 | 1.0<br>7 | 1.3<br>9 |
| **Tested COVID-19^** | 0.01<br>3 | 0.14<br>8 | 0.929 | 1.0<br>1 | 0.7<br>6 | 1.3<br>5 |

\*: $p < 0.05$ ; \*\*: $p < 0.01$ ; \*\*\*: $p < 0.001$ .
^: in a subset of COVID-19-tested individuals (n = 14,796).
SE: standard error; OR: odds ratio; 95% CI (LL; UL): upper and lower limits for the 95% confidence interval of the odds ratio.

The mean age of post-menopausal and pre-menopausal women included in the analysis above was 53.8 and 45.2 years, respectively. Because of this difference, sensitivity analyses were performed for age within 5-year age bins (40–45, 45–50, 50–55 and 55–60 years old). Upon subgroup analysis by age, we observed that *predicted* COVID-19 results were most driven by the 45-50 age group (OR = 1.35, 95% CI 1.05–1.72, *p* = 0.017), wherein anosmia, as well as fever and persistent cough, and the need for oxygen treatment in hospital were also significant (**S2 Table**). A sensitivity analysis was also carried for last period within 3 years at time of reporting. Sensitivity analysis for last menstrual period within 3 years from questionnaire showed similar significant results for higher rate of *predicted* COVID-19 in post-menopausal women (OR = 1.17, 95% CI 1.01–1.36, *p* = 0.036).

### Use of Combined Oral Contraceptive Pill

We examined the link between COCP use and COVID-19-positivity and related outcomes in 295,689 women aged 18–45 years (BMI 20-35). Both pre- and post-menopausal women were considered for this analysis, where most participants (85%) were pre-menopausal. Cases were defined as females on the COCP as their only form of hormonal therapy, resulting in 64,253 COCP-users (**S1 Table)**. Controls were women of the same age and BMI group taking no form of hormone therapy, resulting in 231,436 controls (**S1 Table**).

Women using COCP had a lower rate of *predicted* COVID-19 (OR = 0.87, 95% CI 0.81–0.93, *p* = 8.03 E-05) and a corresponding reduced frequency of symptoms, including persistent cough, delirium, anosmia, skipped meals, severe fatigue and pain (*p* < 0.001) (**Table 2**). Rate of hospitalisation was also significantly lower in COCP users (OR = 0.79, 95% CI 0.64–0.97, *p* = 0.023). The association between COCP use and tested COVID-19 as outcome was not significant but showed a consistent negative direction of association in the 25–30 and 35–40 age groups (**S2 Table**). Analyses in the subset of 251,786 premenopausal women alone, including 52,453 COCP-users and 199,333 controls, showed consistent strong negative associations with *predicted* COVID-19 (OR = 0.87, 95% CI 0.80–0.95, *p* = 8.38E-04) and a corresponding reduced frequency of a wide range of symptoms (**S2 Table**).

**Table 2.**
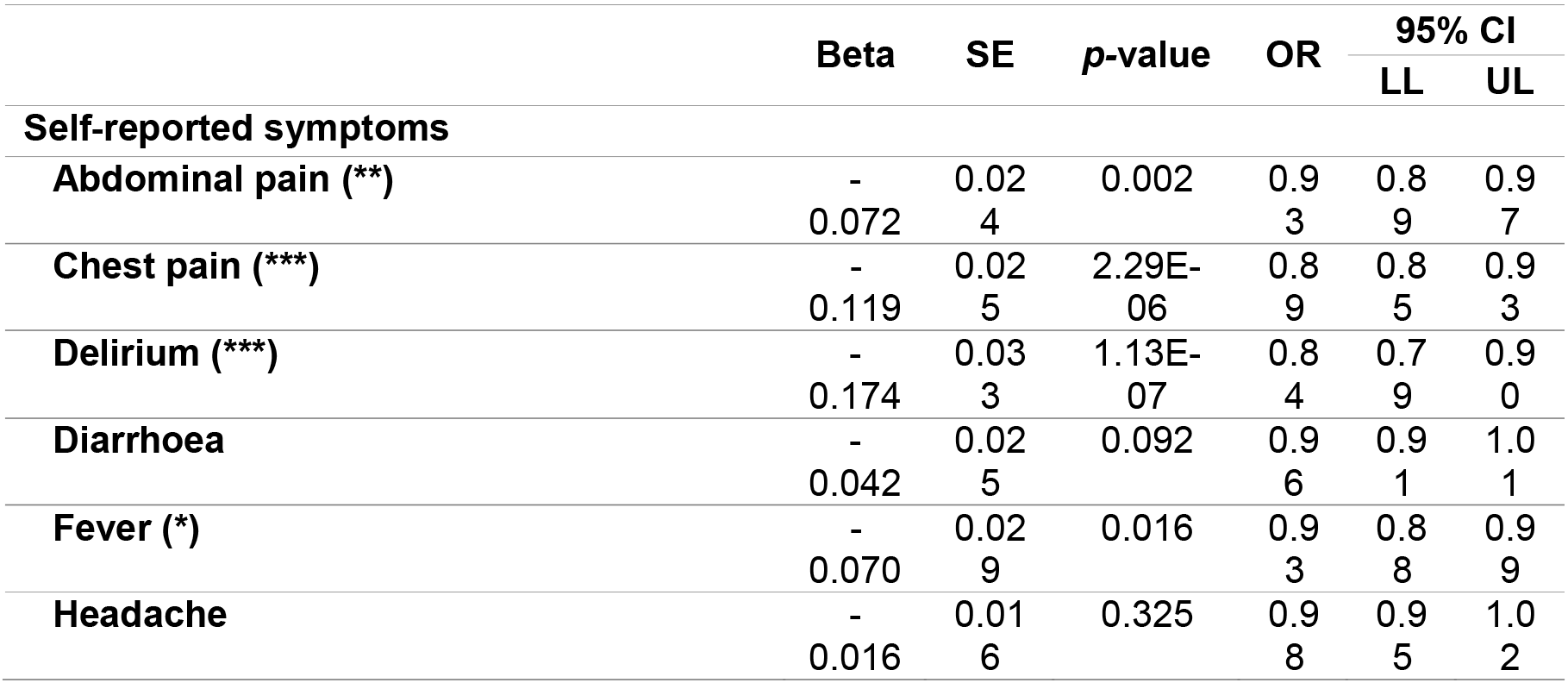
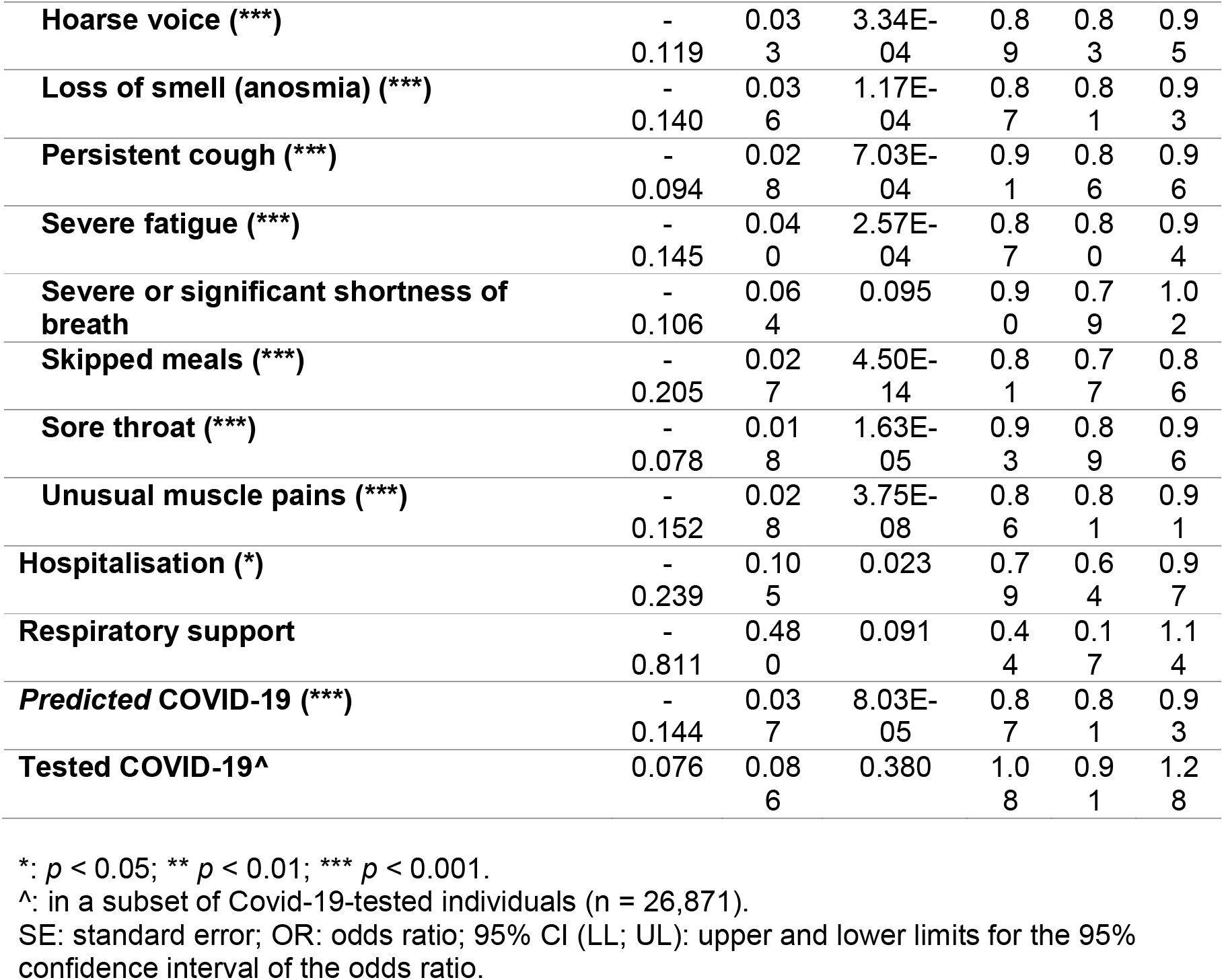
Association of COCP use to *predicted* and tested COVID-19 results, self-reported symptoms, and indicators of COVID-19 disease severity (hospitalisation/respiratory support) (n = 295,689).

The mean ages of COCP users and controls were 29.5 and 34.2 years, respectively. Sensitivity analyses were performed for age, in 5-year bins (18–25, 25–30, 30–35, 35–40 and 40–45-year-olds). Sensitivity analyses showed strongest results for the 25–30 year age group, with COCP-users having lower *predicted* COVID-19-positivity (OR = 0.77, 95% CI 0.67–0.90, *p* = 0.0008) and avoiding hospitalisation (OR = 0.5, 95% CI 0.31–0.80, *p* = 0.004) (**S2 Table**). Negative association with *predicted* COVID-19 was also significant in the 40–45-year-olds (OR = 0.77, 95% CI 0.63–0.94, *p* = 0.01). Associations in the 18–25-year-olds yielded only two statistically significant symptom associations, suggesting that this age group contributed the least to the results observed in the main analysis.

### Use of Hormone Replacement Therapy

The association between HRT use and COVID-19 was assessed in 151,193 post-menopausal women aged 50-65 years, with BMI 20–35 and last periods reported at age 45–60. Controls were post-menopausal women matching these criteria who received no form of hormonal therapy, resulting in 133,395 controls (**S1 Table**). Cases were defined as 17,798 women on HRT only. Extended analyses also considered women on HRT or additional related hormone therapies, excluding use of estrogen in gender transitioning (**S1 Table**).

HRT use was associated with an increased rate of *predicted* COVID-19 (OR = 1.32, 95% CI = 1.16 – 1.49, *p* = 2.22E-05) and frequency of a wide range of symptoms (**S3 Table**). However, while *predicted* COVID-19 and reporting of symptoms showed positive associations in HRT users, there was no significantly increased rate of hospitalisation in HRT-users. Notably, both the need for respiratory support and testing positive for COVID-19 showed a negative trend of association with HRT use, although the results were not nominally significant. The results remained consistent in extended analyses considering use of HRT or other related hormone therapies (**S3 Table**).

The mean ages of HRT users ranged between 56.6 – 56.8 years, while the mean age of controls was 58.2 years. Sensitivity analyses were performed for age, selecting for subgroups of women aged 50–55, 55–60 and 60–65 years. Sensitivity analyses of age were consistent with the overall *predicted* COVID-19 and symptoms results and showed negative directions of association for testing COVID-19-positive, most consistent in 55–60-year-olds (**S2 Table**), which yielded more COVID-19 tests than the 50–55 and 60–65-year-old subgroups (**S1 Table**).

### App data validation in TwinsUK questionnaires

A subset of app users included 270 female research volunteers from the TwinsUK cohort (13). Of these, 84 had previously reported questionnaire data on menopausal status. For all 84 women menopausal status in the app response matched the TwinsUK questionnaire data, where women reported that periods had either stopped or that they did not currently have periods. Furthermore, a subset of 75 female twins had reported information on age of last period in TwinsUK questionnaires. Of these, 64% of twins (48 twins) matched age of last period reported from TwinsUK data within 1 year to age at last period reported in the app, and 87% (65) matched within a 3-year range.

### Menopause, biological aging, and COVID-19 symptoms

Menopause is a marker of ageing and has previously been linked to accelerated epigenetic ageing (23). To this end, we compared the frequency of COVID-19 symptoms in 270 TwinsUK female twins (**S4 Table**) with available app data to 5 estimates of epigenetic ageing in whole blood, including the original epigenetic age acceleration, GrimAge acceleration, PhenoAge acceleration, blood cell *intrinsic* epigenetic age acceleration (IEAA), and blood *extrinsic* epigenetic age acceleration (EEAA).

Overall, fatigue and unusual muscle pains showed the most (3 or more) nominally significant associations with epigenetic age acceleration measures; followed by hoarse voice, skipped meals and anosmia where significant differences were observed for two age acceleration measures; and fever where a significant difference was observed with GrimAge Acceleration alone (p = 0.01) (**S5 Table**). However, the results should be interpreted with caution as sample sizes for subgroup analyses are modest, and in some cases extremely small. Similarly, the number of individuals with *predicted* COVID-19 was extremely small (3 *predicted* cases), but these individuals as a group had on average accelerated epigenetic age acceleration across all five epigenetic ageing measures (**S5 Table**).

## Discussion

Sex is a biological variable that affects immune responses to both self and foreign antigens. The sex of an individual is a multidimensional biological characteristic that shapes infectious disease pathogenesis and is defined by the differential organisation of chromosomes, reproductive organs, and sex steroid levels. Sex is distinct from gender, which includes behaviours and activities that are determined by society or culture in humans. Human biological sex plays a fundamental role in heterogeneous COVID-19 outcomes, with a strong male predominance in mortality. Although gender-related social factors, including smoking, health care-seeking behaviours and some medical comorbidities may impact the outcomes of COVID-19 (24, 25) and contribute to sex-based differences in severity, the cross-cultural emergence of significantly increased risk of mortality for males points to biological risk determinants.

Teasing out the precise drivers of mortality in COVID-19, regardless of sex, is a difficult task. The innate recognition and response to viruses as well as downstream adaptive immune responses during viral infections are known to differ between females and males (26). It has been well-illustrated that females generally mount greater inflammatory, antiviral, and humoral immune responses than males during viral infections (27), which contributes to better clearance of viruses, including SARS-CoV (7). This heightened inflammatory response is advantageous in response to infection and sepsis, but is unfavourable in immune responses against self, leading to an overall increased rate of autoimmune diseases in women compared to men (28, 29). Additionally, enhanced immunity in females can also result in greater immuno-pathology and tissue damage at later stages of viral disease, such as during influenza A virus infection (30). Conversely, maternal physiological adaptations to pregnancy usually predispose pregnant women to a more severe course of many infections, including viral pneumonia, with subsequent higher maternal and fetal morbidity and mortality (31), but observational cohort studies in COVID-19 have reported the risk of severe disease in the pregnant population was approximately half that in the general population of patients presenting with this disease (6). Estrogen levels increase more than 100-fold in pregnancy (32) providing a potential mechanism for the unique resistance to COVID-19 compared with previous pandemic such as H1N1.

With ageing, a general decline in immune function is observed – immune-senescence. Several of these changes are gender specific and affect postmenopausal women. Menopause is a normal part of a woman’s lifecycle and consists of a series of body changes that can last from one to ten years. Levels of estrogen, for example, 17β-estradiol (E2), are variable during the menstrual cycle, high during pregnancy and low after menopause in females. E2 affects many components of innate immunity, including the functional activity of innate immune cells that influence downstream adaptive immune responses (26). Loss of sex hormones due to ageing results in a reduction of immune function. For example, in postmenopausal women, a second peak in Human papilloma virus (HPV) prevalence has been reported (33, 34). New HPV infections in older women with no sexual activity are thought to be due to reduced immune responses (35). HIV-1 infection is also increasing in postmenopausal women (36), where a European study found that women over 45 have a 4-fold increased risk of acquiring HIV compared to women under 45 years of age (37).

As we hypothesised, our results show that being pre-menopausal appears to have a protective effect against COVID-19 in a large community survey of female UK app-users. This was supported by a protective effect seen amongst pre-menopausal women taking the COCP but was not seen for post-menopausal women taking HRT. However, HRT results should be considered with caution due to lack of data on HRT type, route of administration and duration of treatment. Historically, HRT provision and prescribing has been poor in the UK and elsewhere, representing a significant area-of-need in women’s health. There are different effects on various preparations of HRT, and we did not determine the type of HRT that each woman was taking. The bulk of scientific evidence from preclinical, clinical, and epidemiologic studies, and randomised clinical trials clearly indicates that judiciously selected HRT is usually beneficial and rarely dangerous. The majority of women in the UK are currently given oral estrogen, which has more risks compared to transdermal estrogen, and may also affect immunity differently (38). Transdermal estrogen contains E2, which has more beneficial effects on immunity as well as future health (39). Women taking HRT have been shown to have lower future risk of cardiovascular disease, obesity and type 2 diabetes, which are all known to be associated with more severe symptoms and higher mortality from COVID-19 (40).

Menopause is a marker of biological ageing in women that has previously been associated with accelerated epigenetic ageing (21). The associations between menopausal status and COVID-19 positivity and symptom severity may in part be related to biological ageing, rather than reduction in estrogen specifically, although the COCP use results suggest that this is unlikely. To explore the possibility that our results may be attributed to ageing, rather than a reduction in estrogen, we also tested the association between COVID-19 symptoms and epigenetic ageing rates in a subset of participants from the TwinsUK cohorts. Our results are consistent with increased frequency of COVID-19 symptoms among subjects with accelerated biological ageing. However, the sample sizes in the epigenetic subgroup analyses is too modest to draw robust conclusions. Interestingly, despite the extremely small sample size, the group of 3 *predicted* COVID-19 individuals showed significantly greater epigenetic age acceleration, including with epigenetic measures that capture aspects of immune-senescence (EEAA), warranting further investigation into the link between biological ageing and COVID-19 positivity and symptom severity.

This study collected data on hundreds of thousands of women resulting in good power to detect effects. However, our study also has several limitations. Data are self-reported, and questions on medication use were non-standard, to ease large-scale app-based reporting. Data on type, route, duration, and dose of hormone therapies, and importantly HRT, were not collected due to difficulties faced collecting very detailed data using an app-based interface. As such, untangling the effect of differing types of HRT was not possible. Most COCPs contain between 20-35 micrograms of ethinylestradiol along with a progestogen, while HRT estradiol doses are generally lower and more physiological. As such, lower estrogen doses and lack of detailed data may have resulted in the lack of effect seen amongst HRT-users. Another limitation relates to reporting bias within both symptoms and test results. Additionally, sampling using an app will under-represent individuals without smartphones, including older participants, and is likely to under-represent those severely affected by COVID-19. Other limitations include the effects of unmeasured confounding and systematic differences between individuals prescribed different types of hormone therapy. There is also potential for selection bias where, for example, experiencing gynaecological problems and menopause may influence likelihood of hormonal therapy use. To ascertain the consistency of our results, we performed multiple cumulative data extracts over the period 7 May 2020 - 15 June 2020, and observed that association results were consistent throughout, including specifically for *predicted* COVID-19 associations. Nonetheless, because prediction of COVID-19 was based on symptoms reported over a two and half-month period since the app launched, predictions could in some cases be influenced by the reporting of symptoms at far-apart dates. Additionally, the individuals on which the prediction model was trained were highly selected as COVID-19 testing was not performed at random when the app was initially launched, although testing criteria have since been extended and more individuals have been able to access COVID-19 testing.

## Conclusion

Our findings indicate a protective effect of estrogen from symptomatic COVID-19, based on positive association of menopausal status with *predicted* COVID-19, and negative association of COCP use with *predicted* COVID-19. HRT use was positively associated with COVID-19 symptoms. However, the results should be interpreted with caution due to lack of data on HRT type, route of administration, duration of treatment, and potential comorbidities. Further work focussed on gender with hormone profiling in both pre-clinical and clinical settings, as well as on biological ageing, is needed to uncover novel features of the host immune response to SARS-CoV-2 and ultimately result in more equitable health outcomes.

## Data Availability

Data used in this study is available to bona fide researchers through UK Health Data Research using the following link https://healthdatagateway.org/detail/9b604483-9cdc-41b2-b82c-14ee3dd705f6

## Acknowledgments

We thank all participants who entered data into the COVID Symptom Study app, including study volunteers enrolled in the TwinsUK cohort. We thank the staff of the Department of Twin Research at King’s College London and Zoe Global Limited for contributing to the running of the study and data collection.

## Ethics

The App Ethics has been approved by KCL ethics Committee REMAS ID 18210, review reference LRS-19/20-18210 and all subscribers provided consent.

## Declaration of Interests

Zoe Global Limited co-developed the app *pro bono* for non-commercial purposes. Investigators received support from the Wellcome Trust, the MRC/BHF, EU, NIHR, CdRF, and the NIHR-funded BioResource, Clinical Research Facility and BRC based at GSTT NHS Foundation Trust in partnership with KCL. Zoe authors work for Zoe Global Limited and TDS is a consultant to Zoe Global Limited. IB is NIHR Senior Investigator and Chief Data Scientist Advisor for AstraZeneca.

## Funding

This study received no specific funding. However, Zoe Global Limited provided in kind support for all aspects of building, running, and supporting the app and service to all users worldwide. In addition, epigenetic ageing estimates were previously generated with funding from the UK Economic and Social Research Council (ES/N000404/1 to JTB), but the funder had no role in in study design, data collection and analysis, decision to publish, or preparation of the manuscript. Investigators received support from the Wellcome Trust, the MRC/BHF, Alzheimer’s Society, EU, NIHR, CDRF. The TwinsUK study was funded by the Wellcome Trust; European Community’s Seventh Framework Programme (FP7/2007–2013); National Institute for Health Research (NIHR)-funded BioResource, Clinical Research Facility and Biomedical Research Centre based at Guy’s and St Thomas’ NHS Foundation Trust in partnership with King’s College London.

## Contribution Statement

LRN, TDS, CJS and JTB developed the research hypotheses. RC, KAL, LRN and JTB wrote the manuscript. RC performed the primary analyses. BM, MNL, HM, CC, JC-F, JCP, IB, LK, JR and SO contributed to the analyses and interpretation of the results. JCP and JW facilitated the addition of the relevant questions to the app. All authors approved the final manuscript.

Figures

**Supplementary Figure 1.**
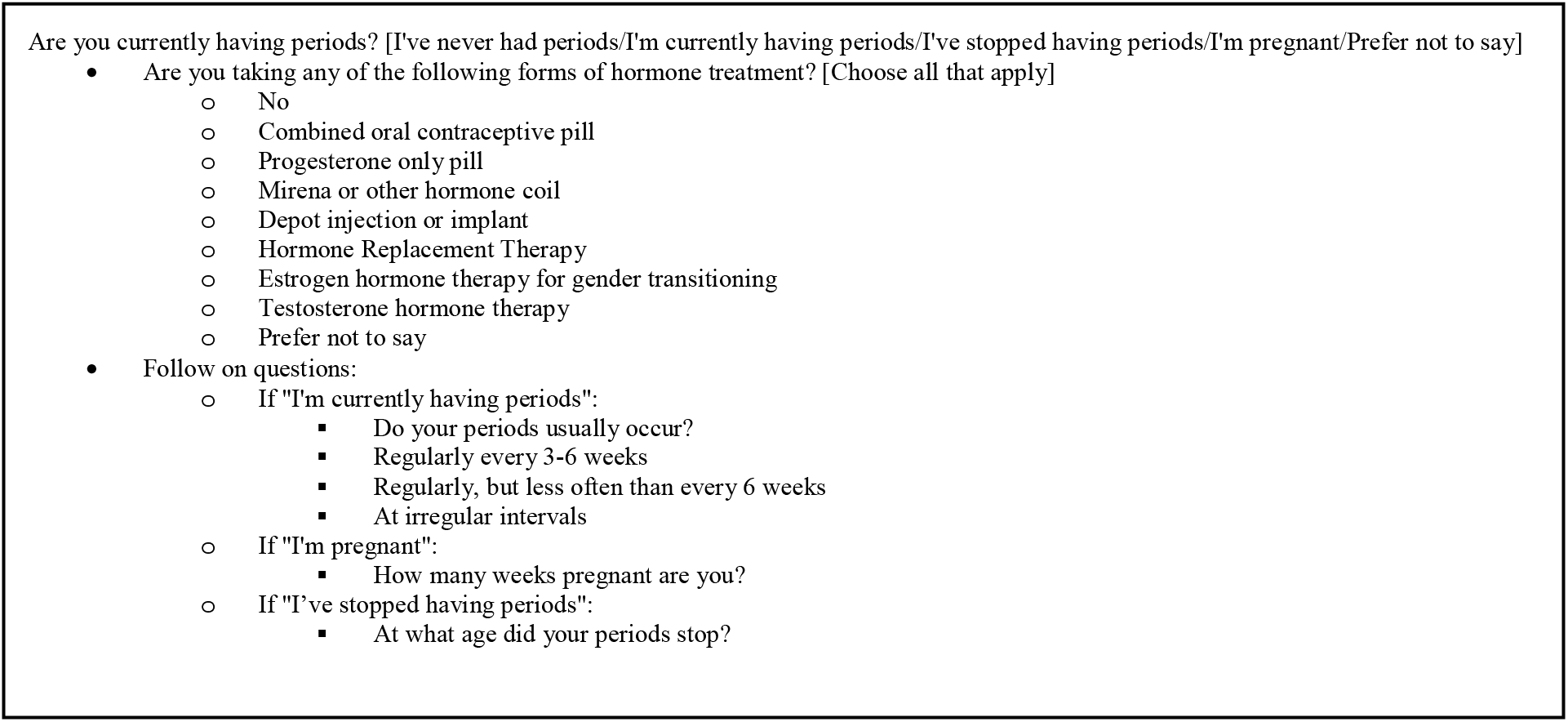
Questions posed to female participants with possible answers

## Notes

### Clinical Trial

The App Ethics has been approved by KCL ethics Committee REMAS ID 18210, review reference LRS-19/20-18210 and all subscribers provided consent. ClinicalTrials.gov Identifier of app: NCT04331509

### Summary of Updates

An author who was on the manuscript hadn't been added to the author list. This has now been done

